# Real-Time SARS-CoV-2 Genotyping by High-Throughput Multiplex PCR Reveals the Epidemiology of the Variants of Concern in Qatar

**DOI:** 10.1101/2021.07.18.21260718

**Authors:** Mohammad R. Hasan, Mahesh K. R. Kalikiri, Faheem Mirza, Sathyavathi Sundararaju, Anju Sharma, Stephan Lorenz, Hiam Chemaitelly, Reham A. El-Kahlout, Kin Ming Tsui, Hadi M. Yassine, Peter V. Coyle, Abdullatif Al Khal, Roberto Bertollini, Mohamed H. Al Thani, Laith J. Abu-Raddad, Patrick Tang, National Study Group for COVID-19 Epidemiology in Qatar

## Abstract

Complementing whole genome sequencing strategies with high-throughput multiplex RT-qPCR genotyping allows for more comprehensive and real-time tracking of SARS-CoV-2 variants of concern. During the second and third waves of COVID-19 in Qatar, PCR genotyping, combined with Sanger sequencing of un-typeable samples, was employed to describe the epidemiology of the Alpha, Beta and Delta variants. A total of 9792 nasopharyngeal PCR-positive samples collected between April-June 2021 were successfully genotyped, revealing the importation and transmission dynamics of these three variants in Qatar.

---

The emergence of severe acute respiratory syndrome coronavirus-2 (SARS-CoV-2) variants of concern (VOC) with higher transmissibility, increased virulence and/or ability to escape natural or vaccine-derived immunity has threatened the extraordinary public health and vaccination efforts against coronavirus disease 2019 (COVID-19). These SARS-CoV-2 variants, which have spread to many countries around the world, include Alpha, Beta, Gamma and Delta (Pango lineage B.1.1.7, B.1.351, P.1 and B.1.617.2, respectively) VOCs (1). Therefore, in addition to the routine surveillance for SARS-CoV-2, it is important to monitor the epidemiology of these VOCs to determine the effectiveness of vaccines (2, 3) and public health measures to prevent their spread.

SARS-CoV-2 variant typing and lineage assignment are typically performed by sequencing of the entire viral genome or the spike gene by next-generation sequencing (NGS). Although whole genome sequencing provides the most complete data for genetic characterization and monitoring the evolution of the virus, it is expensive and may not be fast enough for timely public health responses. Furthermore, many countries may lack the infrastructure and resources needed for large-scale screening of SARS-CoV-2 variants through next-generation sequencing. Alternatively, Sanger sequencing of a targeted region of the genome can be both informative and less costly, but it is time consuming and less amenable to high-throughput (4). To this end, multiplex RT-qPCR for differentiating the major variants may serve as an important complementary approach for large-scale screening for SARS-CoV-2 VOCs.

We employed a previously described multiplex RT-qPCR genotyping strategy targeting the Δ69/70 HV deletion in the spike gene and the Δ3675-3677 SGF deletion in the ORF1a gene to distinguish the Alpha variant from Beta/Gamma variants (5). The method also includes CDC N1 assay to identify ‘other’ non-Alpha and non-Beta/Gamma variants for further subtyping. To characterize the ‘other’ category, we performed Sanger sequencing of the spike gene receptor binding domain (RBD). With the aid of liquid handling robotics and with minor modifications to the method, we have adapted the previously described strategy to scale-up variant screening capacity and to generate real-time, actionable epidemiological data on SARS-CoV-2 VOCs in Qatar.

## Verification of SARS-CoV-2 variant PCR assay against Sanger sequencing of the RBD

Variant PCR was performed on 33 nasopharyngeal swab (NPS) specimens positive for SARS-CoV-2 as tested by the Xpert Xpress SARS-CoV-2 assay (Cepheid), and variant calls were made with minor modifications to the original method (Supplemental methods). All samples also underwent Sanger sequencing of the receptor binding domain (RBD) of surface glycoprotein (S) gene to detect the N501Y, E484K, L452R and T478K mutations. SARS-CoV-2 with only N501Y mutation was designated as B.1.1.7-like and those harboring both N501Y and E484K mutations were designated as B.1.351/P.1-like. All other genotypes were designated as ‘other’. With reference to Sanger sequencing, variant PCR correctly predicted 32 out of 33 results (Supplemental table 3). One sample was falsely genotyped as B.1.1.7 by variant PCR while RBD sequencing was consistent with B.1.351/P.1.

## Scaling up SARS-CoV-2 variant PCR for large-scale screening

Between March 27, 2021 and June 19, 2021, a total of 13,797 randomly selected, SARS-CoV-2 positive NPS specimens were received on a weekly basis from the national COVID-19 biorepository at the Qatar Biobank. The number of samples received ranged from approximately 10% to 47% of the total number of new positive cases reported in the country each week (Figure 1A) (https://www.data.gov.qa/pages/home/). All samples were inactivated with a lysis buffer, de-identified and assigned with a barcoded, sample identifier prior to transport to the laboratory at Sidra Medicine, where PCR genotyping was performed. Decoding of barcodes, sample transfer for extraction, and de-capping and re-capping of storage vials were automated, using a liquid handler (Hamilton Germany GmbH). Viral RNA was extracted in a KingFisher Flex system (Thermofisher Scientific) and multiplex, RT-qPCR for SARS-CoV-2 VOCs was performed in batches of 96-samples in an ABI7500 system (Thermofisher Scientific)(Supplemental methods). In this setting, an approximate capacity of extraction and variant PCR for 1000 samples per day was achieved. Variant calls were made based on the differential of the RT-qPCR Ct values for the N gene, and Δ69/70 HV and Δ3675-3677 SGF deletions (Supplemental methods).

**Figure 1:**
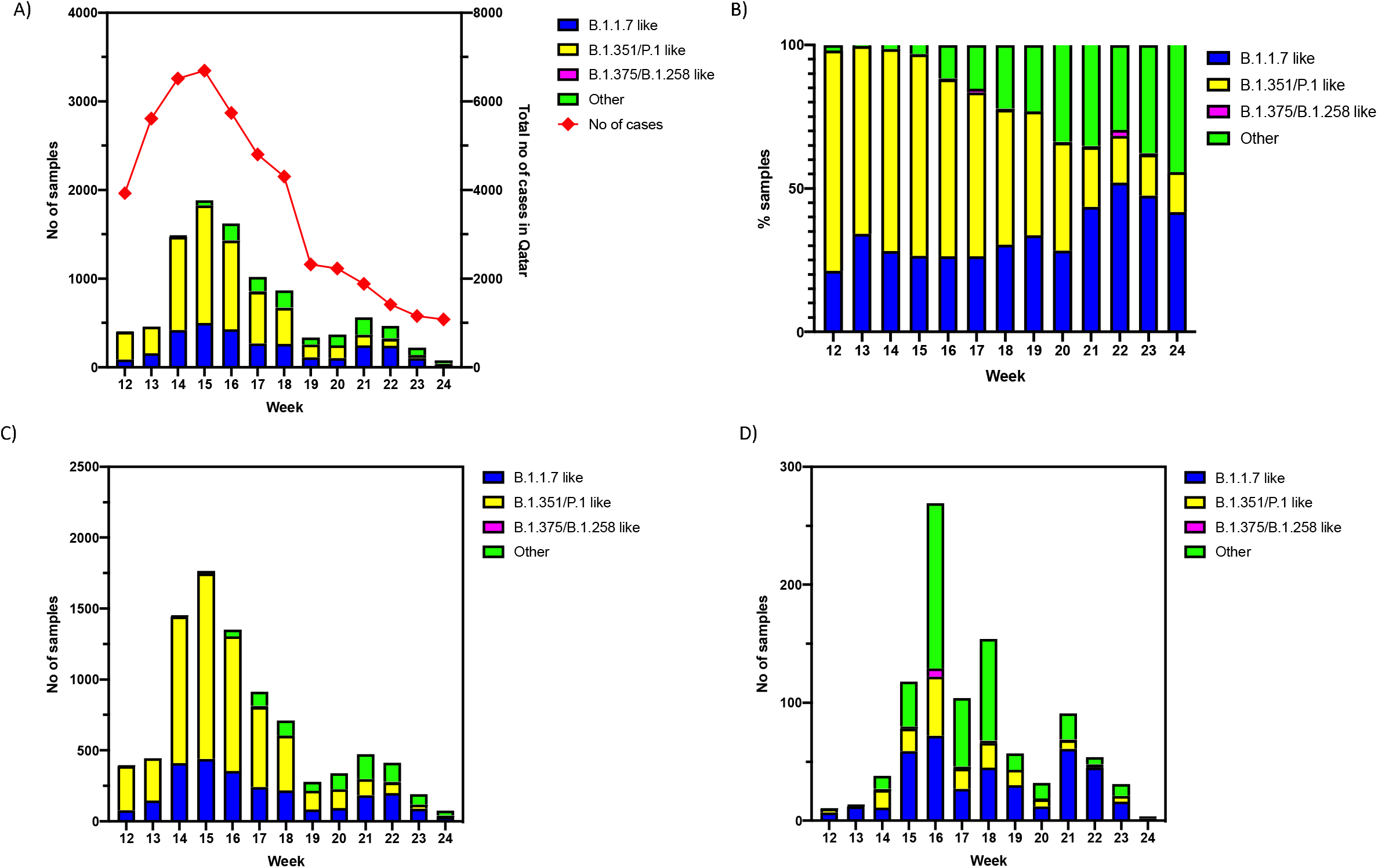
Transmission dynamics of SARS-CoV-2 VOCs in Qatar during April – June 2021. A) Number of cases of SARS-CoV-2 VOCs in the overall population by week of swab collection B) Proportion (%) of cases of SARS-CoV-2 VOCs in the overall population by week of swab collection C) Number of cases of SARS-CoV-2 VOCs in the community by week of swab collection D) Imported cases of SARS-CoV-2 VOCs by week of swab collection.

A total of 9792 samples were successfully genotyped by the variant PCR, and 4005 samples remained undetermined because of weak or undetermined RT-qPCR Ct values. The proportion of samples that remained undetermined by the variant PCR remained similar (∼30%) each week except for week 22 (Supplemental figure 1) when a higher proportion of samples were successfully genotyped. Overall, 2970 (30.3%) were B.1.1.7-like, 5500 (56.2%) were B.1.351/P.1-like, 40 (0.4%) were B.1.375/B.1.258-like and 1282 (13.1%) were ‘other’ variants. From the ‘other’ category, 109 were subjected to Sanger sequencing and 103 (94.5%) were B.1.617.2-like, 3 were B.1-like (2.8%) and 3 (2.8%) were undetermined. Apart from these variants, 56 samples from the other category gave negative results for the N gene but were positive for one or more of the other targets. These variants likely represent the N-gene variants we reported earlier (6).

## Epidemiology of SARS-CoV-2 VOCs in Qatar based on variant PCR

In order to describe the epidemiology of the VOCs, the collection date, age, sex, nationality, reason for testing, and vaccination status associated with each sample were extracted from a federated, national SARS-CoV-2 database. The second and third waves of the COVID-19 pandemic in Qatar were predominantly driven by the spread of the B.1.1.7 and B.1.351 variants, respectively (2, 3). The large third wave, dominated by the B.1.351 variant (2, 3), led to the large contribution of B.1.351/P.1-like cases in the multiplex PCR testing. As the epidemic began to decline in Qatar, a large number of cases with ‘other’ variants were detected, coincident with the epidemics in India and Nepal (Figure 1A and 1B), where a third of the resident population of Qatar originates from (7, 8). Of note, Qatar has diverse demographics where 89% of the population are expatriates from over 150 countries (7, 8).

Although the cases with B.1.351/P.1-like variants dominated both in number and proportion during the third wave between March-May, the proportion of B.1.1.7-like and other variants increased significantly as the epidemic declined following this wave. The epidemiology of the SARS-CoV-2 VOCs in the community was similar to the overall epidemiology (Figure 1C). However, a large number of B.1.1.7–like and ‘other’ variants were detected from the imported cases coincident with the outbreaks in India and Nepal (Figure 1D) and the importation of the ‘other’ variant declined significantly after targeted travel restrictions were implemented during week 17. At the port of entry, the ‘other’ variants were predominantly detected in individuals with Indian and Nepalese nationality (Figure 2A). In the community, the ‘other’ variant was initially seen within those with Indian nationality, but it eventually spread to other nationalities.

**Figure 2:**
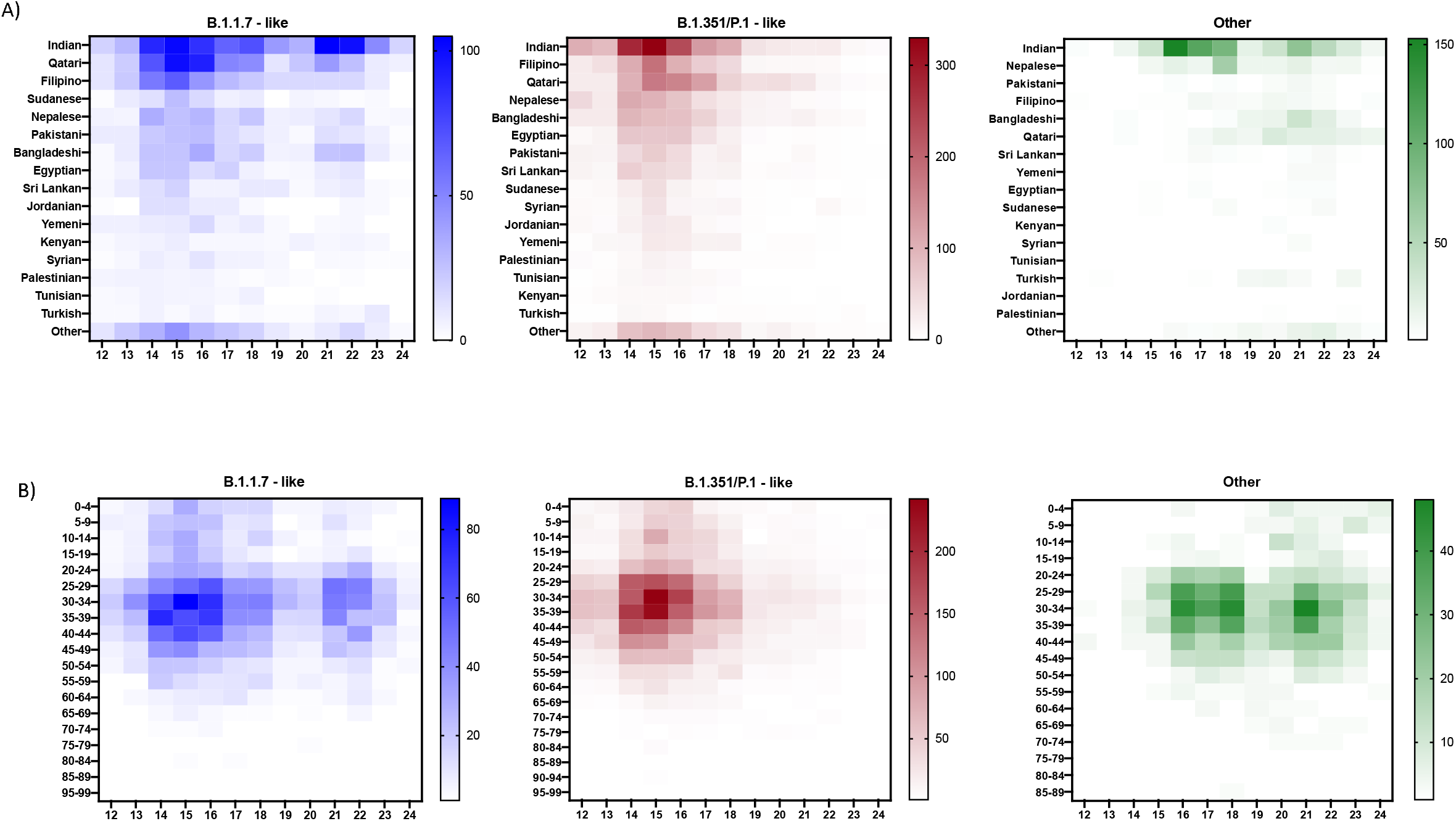
Distribution of cases with SARS-CoV-2 VOCs. A) Number of cases of SARS-CoV-2 VOCs by nationality B) Number of cases of SARS-CoV-2 VOCs by age

**Figure 3:**
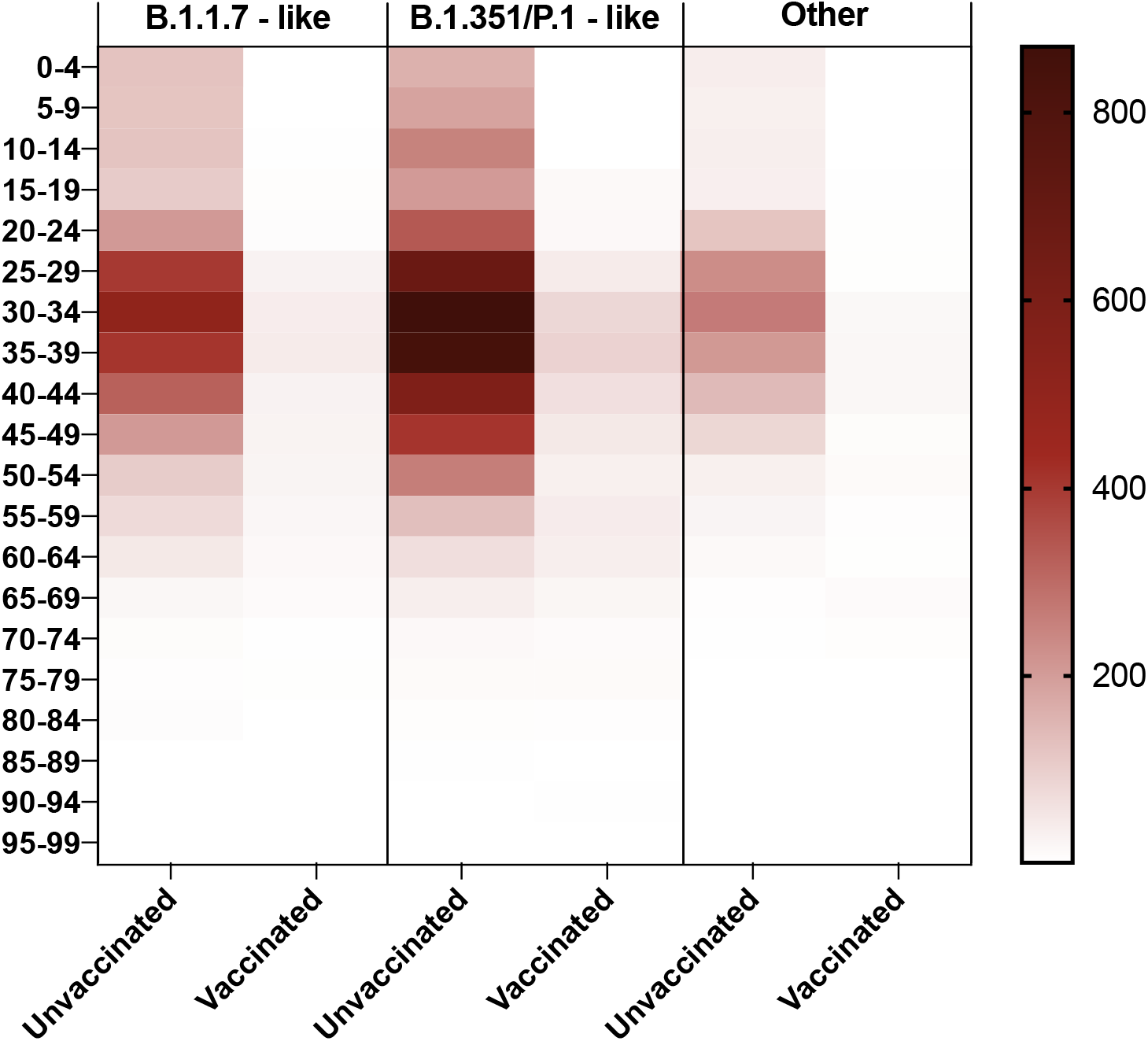
Number of cases of SARS-CoV-2 VOCs by vaccine status.

Secondary small peaks for B.1.1.7-like and ‘other’ variants were noted in weeks 21 and 22 reflecting probably community transmission after their importation (Figure 2A and 2B). For each variant, most cases during the second and third waves were in individuals between 25-44 years old and in those who were not fully vaccinated. However, a larger number of vaccinated individuals were infected by B.1.351/P.1-like variants compared to the other variants, as a consequence of the lower protection for the BNT162b2 vaccine against the B.1.351 variant (2).

## Discussion

We have applied a multiplex RT-qPCR adapted for rapid and large-scale genotyping of SARS-CoV-2 VOCs to describe their real-time epidemiology within Qatar. While the variant PCR was designed to detect B.1.1.7-like and B.1.351/P.1-like variants, RBD sequencing of the ‘other’ type revealed that most variants within this category were B.1.617.2-like. The Alpha and Beta variants were responsible for the large second and third waves of COVID-19 infections in Qatar, respectively. As the third wave declined, a large importation of Delta variant cases occurred, consistent with the timing of the large epidemic in India and the large Indian population residing in Qatar (7, 8). However, despite the influx of the Delta variant into the community, it was unable to generate a significant outbreak. This is likely due to the high level of immunity in the community from the mass vaccination campaign (2, 3) and previous infection (9-13), as well as the continuation of various public health measures against the spread of SARS-CoV-2, including quarantine requirements for returning travelers, and active surveillance efforts including those described in this study. With the decline of the epidemic in Qatar to a low incidence phase, the resulting distribution of the three dominant variants (Alpha, Beta, and Delta) suggests higher ‘effective reproduction number or transmissibility of Alpha and Delta variants that makes them more successful versus the Beta variant (14), despite the greater immune evasion capacity of the Beta variant (15, 16).

In conclusion, we have demonstrated that actionable epidemiological data can be generated by variant PCR so that important public health decisions can be taken in a timely manner. While WGS is still necessary to track the evolution of the virus over time in a given country, most public health decisions do not require such high-resolution data. In particular, for countries lacking the technical capability and resources for large-scale surveillance based on WGS, variant PCR may be a useful tool in informing national COVID-19 responses and preventing the importation of SARS-CoV-2 VOCs and reducing their spread in the community.

## Data Availability

Representative SARS-CoV-2 sequence data generated in this study have been deposited in GeneBank with the accession codes MZ571211-MZ571216.

https://www.ncbi.nlm.nih.gov

