## Supplemental information for "Real-Time SARS-CoV-2 Genotyping by High-Throughput Multiplex PCR Reveals the Epidemiology of the Variants of Concern in Qatar"

**Supplemental methods:**

**Sample collection:** The SARS-CoV-2 positive samples collected in universal transport medium (UTM) were mixed with an equal volume of NUCLISENS easyMAG lysis buffer (BioMerieux) to lyse cells and inactivate the virus. 0.4 ml of inactivated specimens were transferred to 0.5 ml FluidX External Thread Jacket, 2D barcoded tubes in 96-well plate format and were stored at -20°C until tested.

**Variant PCR:** FluidX plates were thawed at 4°C and vortexed for 3 min at 1800 rpm on a plate shaker (Digital MicroPlate Genie Pulse, Scientific Industries Inc. USA). Plates were centrifuged at 2000 rpm for 30 sec (Eppendorf 5804/ 5804 R - Benchtop Centrifuge) immediately before de-capping. FluidX plate and 0.5 ml tube 2D barcodes were decoded, and the tubes were de-capped/re-capped using a Hamilton LabElite DeCapper integrated to a Hamilton Microlab STAR NGS (Hamilton AG). 0.2 ml of specimen was transferred from FluidX tubes to KingFisher™ Deep Well 96 plate. Viral RNA was extracted in batches of 96 samples using the MagMAX Viral and Pathogen Nucleic Acid Isolation Kit in a KingFisher Flex system (Thermofisher Scientific) according to the manufacturer's method. PCR set-up was simplified and expedited by automated dispensing of a reaction mix containing primers and probes for the variant assay and TaqPath™ 1-Step RT-qPCR Master Mix (Thermofisher Scientific) and the transfer of specimens with the aid of multichannel pipettes. Sample identification numbers were imported to ABI7500 fast software and real-time PCR was performed in an ABI7500 system (Thermofisher Scientific).

For samples that were positive for all 3 PCR targets (CDC_N1, Yale_69/70del and Yale_Orf1a-del), mean (±SD) Ct value difference of Yale_69/70del and Yale_Orf1a-del from CDC_N1 were -0.42±1.3 and 0.002±1.6, respectively, and median (IQR) Ct value difference of Yale_69/70del and Yale_Orf1a-del from CDC_N1 were -0.38 (-0.76 to 0.02) and 0.23 (-0.31 to 0.64), respectively. Variant calls were made as described previously (reference 3) except that RT-qPCR Ct value differences of -6.6 (100-fold) were considered and interpreted as shown in supplemental table 1.

**Sequencing:** For Sanger sequencing, the N-terminal and receptor-binding (RBD) regions of SARS-CoV-2 surface glycoprotein (S) gene were amplified with primer set 1 (1S_sarb-Seq-F1: AGGGGTACTGCTGTTATGTCT and 1S_sarb-Seq-R1: CAGTGGAAGCAAAATAAACACCA) and primer set 2 (4S_sarb-Seq-F1: AGATGATTTTACAGGCTGCGT and 4S_sarb-Seq-R1: ACACTGACACCACCAAAAGA), respectively, using OneStep RT-PCR Kit (Qiagen). The concentration of each primer in the final reaction was 0.2 μM. Sequencing was performed, and sequence data were analyzed as described previously (Reference 6). Variant calls were made according to supplemental table 2.

**Supplemental Table 1: Interpretation of variant PCR results**

| CDC_N1 (FAM) | Yale_69/70del (HEX) | Yale_Orf1a-del (Cy5) | Result |
|---|---|---|---|
| CT ≤ 35 | Undetermined or $^{1}\Delta$Ct ≤ -6.6 | Undetermined | B.1.1.7-like |
| CT ≤ 35 | CT ≤ 35 and $^{1}\Delta$Ct ≥ -6.6 | Undetermined or $^{2}\Delta$Ct ≤ -6.6 | B.1.351/P.1-like |
| CT ≤ 35 | Undetermined or $^{1}\Delta$Ct ≤ -6.6 | CT ≤ 35 and $^{2}\Delta$Ct ≥ -6.6 | B.1.375/B.1.258-like |
| CT ≤ 35 | CT ≤ 35 and $^{1}\Delta$Ct ≥ -6.6 | CT ≤ 35 and $^{2}\Delta$Ct ≥ -6.6 | Other |
| CT > 35 or Undetermined | CT ≤ 35 or $^{1}\Delta$Ct ≤ -6.6 | CT ≤ 35 or $^{2}\Delta$Ct ≤ -6.6 | Other- potential N gene variant |
| CT > 35 or Undetermined | CT > 35 or Undetermined | CT > 35 or Undetermined | Undetermined |

$^{1}\Delta$Ct = (Ct value of CDC_N1 - Ct value of Yale_69/70del); $^{2}\Delta$Ct = (Ct value of CDC_N1 - Ct value of Yale_Orf1a-del)

**Supplemental Table 2: Interpretation of sequencing data**

| N-terminal domain | | RBD domain | | | | Interpretation |
|---|---|---|---|---|---|---|
| T19R | delta69 | L452R | T478K | E484K | N501Y | |
| - | - | ND | ND | ND | ND | B.1-like |
| ND | ND | - | - | - | - | B.1-like |
| ND | ND | - | - | - | √ | B.1.1.7-like |
| ND | ND | - | - | √ | √ | B.1.351-like |
| ND | ND | √ | √ | - | - | B.1.617.2-like |
| √ | - | ND | ND | ND | ND | B.1.617.2/B.1.617.3-like |

ND = not done; √ = mutation detected; “-“ = no mutation detected

**Supplemental Table 3: Comparison of SARS-CoV-2 variant PCR results with the results from Sanger sequencing of Spike protein Receptor Binding Domain (RBD)**

| Sample No | Multiplex RT-qPCR | | | | Sanger sequencing | | | | |
|---|---|---|---|---|---|---|---|---|---|
| | NcOv-n1 | YALE-69 | YALE-ORF | PCR result | L452R | T478K | E484K | N501Y | Sanger result |
| 1 | 19.5 | 20.0 | Undetermined | B.1.351/P.1 like | No | No | Yes | Yes | B.1.351/P.1 like |
| 2 | 21.8 | Undetermined | Undetermined | B.1.1.7 like | No | No | No | Yes | B.1.1.7 like |
| 3 | 21.6 | Undetermined | Undetermined | B.1.1.7 like | No | No | No | Yes | B.1.1.7 like |
| 4 | 24.4 | 24.8 | Undetermined | B.1.351/P.1 like | No | No | Yes | Yes | B.1.351/P.1 like |
| 5 | 24.2 | 24.4 | Undetermined | B.1.351/P.1 like | No | No | Yes | Yes | B.1.351/P.1 like |
| 6 | 23.6 | 23.7 | Undetermined | B.1.351/P.1 like | No | No | Yes | Yes | B.1.351/P.1 like |
| 7 | 17.5 | 17.8 | Undetermined | B.1.351/P.1 like | No | No | Yes | Yes | B.1.351/P.1 like |
| 8 | 20.7 | 36.3 | Undetermined | B.1.1.7 like | No | No | No | Yes | B.1.1.7 like |
| 9 | 18.7 | 19.1 | Undetermined | B.1.351/P.1 like | No | No | Yes | Yes | B.1.351/P.1 like |
| 10 | 17.1 | 27.8 | Undetermined | B.1.351/P.1 like | No | No | Yes | Yes | B.1.351/P.1 like |
| 11 | 24.7 | 24.9 | Undetermined | B.1.351/P.1 like | No | No | Yes | Yes | B.1.351/P.1 like |
| 12 | 18.7 | 19.0 | Undetermined | B.1.351/P.1 like | No | No | Yes | Yes | B.1.351/P.1 like |
| 13 | 16.7 | 28.1 | Undetermined | B.1.351/P.1 like | No | No | No | Yes | B.1.1.7 like |
| 14 | 17.9 | 18.2 | Undetermined | B.1.351/P.1 like | No | No | Yes | Yes | B.1.351/P.1 like |
| 15 | 21.3 | 21.7 | Undetermined | B.1.351/P.1 like | No | No | Yes | Yes | B.1.351/P.1 like |
| 16 | 24.7 | 25.4 | Undetermined | B.1.351/P.1 like | No | No | Yes | Yes | B.1.351/P.1 like |
| 17 | 17.6 | 19.9 | 19.2 | Other | No | No | No | No | B.1 like |
| 18 | 19.7 | 28.7 | Undetermined | B.1.1.7 like | No | No | No | Yes | B.1.1.7 like |
| 19 | 28.2 | Undetermined | Undetermined | B.1.1.7 like | No | No | Yes | Yes | B.1.351/P.1 like |
| 20 | 15.9 | 16.3 | Undetermined | B.1.351/P.1 like | No | No | Yes | Yes | B.1.351/P.1 like |
| 21 | 16.0 | 16.4 | Undetermined | B.1.351/P.1 like | No | No | Yes | Yes | B.1.351/P.1 like |
| 22 | 22.9 | 23.2 | Undetermined | B.1.351/P.1 like | No | No | Yes | Yes | B.1.351/P.1 like |
| 23 | 26.7 | 28.4 | 29.1 | Other | No | No | No | No | B.1 like |
| 24 | 22.2 | 24.3 | 23.8 | Other | No | No | No | No | B.1 like |
| 25 | 23.7 | 23.3 | 24.0 | Other | No | No | No | No | B.1 like |
| 26 | 16.6 | Undetermined | Undetermined | B.1.351/P.1 like | No | No | Yes | Yes | B.1.351/P.1 like |
| 27 | 20.1 | Undetermined | Undetermined | B.1.351/P.1 like | No | No | Yes | Yes | B.1.351/P.1 like |

| | | | | | | | | | |
|---|---|---|---|---|---|---|---|---|---|
| 28 | 18.6 | Undetermined | Undetermined | B.1.351/P.1 like | No | No | Yes | Yes | B.1.351/P.1 like |
| 29 | 18.8 | Undetermined | Undetermined | B.1.351/P.1 like | No | No | Yes | Yes | B.1.351/P.1 like |
| 30 | 16.6 | 16.9 | Undetermined | B.1.1.7 like | No | No | No | Yes | B.1.1.7 like |
| 31 | 14.8 | 16.0 | Undetermined | B.1.1.7 like | No | No | No | Yes | B.1.1.7 like |
| 32 | 12.8 | 13.7 | Undetermined | B.1.1.7 like | No | No | No | Yes | B.1.1.7 like |
| 33 | 16.7 | 17.8 | Undetermined | B.1.1.7 like | No | No | No | Yes | B.1.1.7 like |

**National Study Group for COVID-19 Epidemiology in Qatar**

Laith J. Abu Raddad, Ph.D.
Hiam Chemaitelly, M.Sc.
Joel A. Malek, Ph.D.
Weill Cornell Medicine-Qatar, Cornell University, Doha, Qatar

Fatiha M. Benslimane, PhD
Hebah A. Al Khatib, PhD
Hadi M. Yassine, Ph.D.
Houssein H. Ayoub, Ph.D.
Hanan F. Abdul Rahim, Ph.D.
Gheyath K. Nasrallah, Ph.D.
Qatar University, Doha, Qatar

Adeel A. Butt, M.D. M.S.
Peter Coyle, M.D.
Andrew Jeremijenko, MD
Zaina Al Kanaani, Ph.D.
Abdullatif Al Khal, M.D.
Einas Al Kuwari, M.D.
Anvar H. Kaleeckal, M.Sc.
Ali Nizar Latif, M.D.
Riyazuddin M. Shaik, M.Sc.
Hamad Medical Corporation, Doha, Qatar

Patrick Tang, M.D. Ph.D.
Mohammad R. Hasan, Ph.D.
Sidra Medicine, Doha, Qatar

Mohamed Ghaith Al Kuwari, M.D.,

Primary Health Care Corporation, Doha, Qatar

Roberto Bertollini, M.D., M.P.H.
Hamad Eid Al Romaihi, M.D.
Mohamed H. Al Thani, M.D., M.P.H.
Ministry of Public Health, Doha, Qatar
